# Breast imaging with an ultra-low field MRI scanner: a pilot study

**DOI:** 10.1101/2024.04.01.24305081

**Authors:** Sheng Shen, Neha Koonjoo, Friderike K. Longarino, Leslie R. Lamb, Juan C. Villa Camacho, Torben P.P. Hornung, Stephen E. Ogier, Susu Yan, Thomas R. Bortfeld, Mansi A. Saksena, Kathryn E. Keenan, Matthew S. Rosen

**Affiliations:** A.A. Martinos Center for Biomedical Imaging, Department of Radiology, Massachusetts General Hospital, Boston, MA, USA; Harvard Medical School, Boston, MA, USA; Department of Radiation Oncology, Radiation Biophysics Division, Massachusetts General Hospital, Boston, MA, USA; Clinical Cooperation Unit Translational Radiation Oncology, German Cancer Research Center (DKFZ), Heidelberg, Germany; Department of Radiation Oncology, Heidelberg University Hospital, Heidelberg, Germany; Massachusetts General Hospital, Division of Breast Imaging, Boston, MA, USA; Department of Physics, ETH Zürich, Zürich, Switzerland; Physical Measurement Laboratory, National Institute of Standards and Technology, Boulder, CO, USA; Department of Physics, University of Colorado, Boulder, CO, USA; Department of Physics, Harvard University, Cambridge, MA, USA

## Abstract

Breast cancer screening is necessary to reduce mortality due to undetected breast cancer. Current methods have limitations, and as a result many women forego regular screening. Magnetic resonance imaging (MRI) can overcome most of these limitations, but access to conventional MRI is not widely available for routine annual screening. Here, we used an MRI scanner operating at ultra-low field (ULF) to image the left breasts of 11 women (mean age, 35 years ±13 years) in the prone position. Three breast radiologists reviewed the imaging and were able to discern the breast outline and distinguish fibroglandular tissue (FGT) from intramammary adipose tissue. Additionally, the expert readers agreed on their assessment of the breast tissue pattern including fatty, scattered FGT, heterogeneous FGT, and extreme FGT. This preliminary work demonstrates that ULF breast MRI is feasible and may be a potential option for comfortable, widely deployable, and low-cost breast cancer diagnosis and screening.

## Introduction

Approximately 1 in 8 women will develop breast cancer in their lifetime (*1*), with 85% of cancers occurring in women with no family history of breast cancer (*2*). Currently mammography is the most used imaging-based tool for breast cancer screening as it is accessible and cost-effective. However, mammography has limitations: it requires ionizing radiation, women find breast compression uncomfortable, and 1-35% of breast cancers are missed on mammograms (*3–9*). As a result, in 2015, only 65.3% of women over age 40 had undergone a mammogram in the previous 2 years (*10*).

Currently available MRI-based methods overcome some of these limitations (*11*), particularly in high-risk groups (*12–14*). This is because differences in soft tissues can be visualized without obfuscations from dense tissue, and MRI screening has low false-negative rates (*15, 16*). MRI can detect invasive carcinomas, distinguishing between malignant and benign lesions using T1 weighted imaging with injected contrast agent enhancement (*17*). Additionally, apparent diffusion coefficient (ADC) can be used to differentiate lesions (*18*) and assess response to treatment(*19*). However, traditional clinical MRI operating at 1.5 T and 3 T requires the patient to endure a constricted setting, and currently, MRI as a screening modality is underutilized in high-risk women (defined as a lifetime risk >20%) (*20*). While fast MRI protocols enable screening in less than 10 minutes (*21*), the high cost and limited access prohibit their use as a primary screening tool.

Compared to clinical MRI systems operating at 1.5 T or 3 T, ultra-low field (ULF, <10 mT) MRI systems can be significantly less expensive to build and have less stringent installation requirements, allowing increased access. Recently, low-field MRI neuroimaging systems operating at 64 mT have been used in the clinic at the patient bedside for stroke detection (*22–24*) These systems are safe, do not require an MRI technician, do not require a magnetic- or RF-shielded room, and can be rolled from room to room (*25, 26*). While operation at lower magnetic field generally leads to images obtained with lower SNR, the effectiveness of low-field MRI for neuroimaging in clinical practice has been demonstrated (*22, 24, 27*).

Based on the recent successes of low field MRI for neuroimaging, we hypothesize that there may be sufficient SNR for whole breast imaging with ULF MRI. NMR-based methods to assess breast cancers began in the early days of MRI with work at 0.71 T to measure T1 and T2 relaxation times of breast tissues (*28*) Given those promising results, T1 was measured on an entire mastectomy sample (*29, 30*) and then the whole breast was imaged at 45 mT, which supported the NMR findings, although adoption of the method was limited by an unacceptably long exam duration (*31*). Other studies report the T1 relaxation times of *ex vivo* breast tissues at a range of magnetic fields using NMR dispersion (NMRD) measurements (*32, 33*). These works found that in the low- and ultra-low field regime the T1 relaxation time of cancerous breast lesions differs from that of healthy fibroglandular and adipose tissues (*28, 32, 33*). These T1 differences motivate the presently described work; if one could obtain sufficient SNR over a reasonable exam time, ULF breast MRI may be suitable for low-cost breast imaging, retaining the advantages of multi-slice soft tissue imaging compared to the X-ray projection-based method used in mammography.

In this study, we describe our preliminary evaluation of breast imaging using ULF MRI. We used an ULF MRI system operating at 6.5 mT and a conical RF coil to image the left breasts of 11 women in the same prone position. ULF MR images of the whole breast revealed essential breast features, including type of fibroglandular tissue, breast outline, nipple areolar complex, and chest wall. These findings are encouraging, and ULF breast MRI may indeed be suitable as a strategy to increase access for comfortable, non-invasive breast imaging.

## Results

### Imaging system

Imaging was performed on a custom-built electromagnet-based MRI scanner shown in Fig. 1 and modified for breast imaging from its previously described configuration for neuroimaging (*34*). Figure 2 shows the imaging bed and dedicated RF coil designed to image a single breast. The breast and breast RF coil are placed at the isocenter of the scanner.

**Fig. 1.**
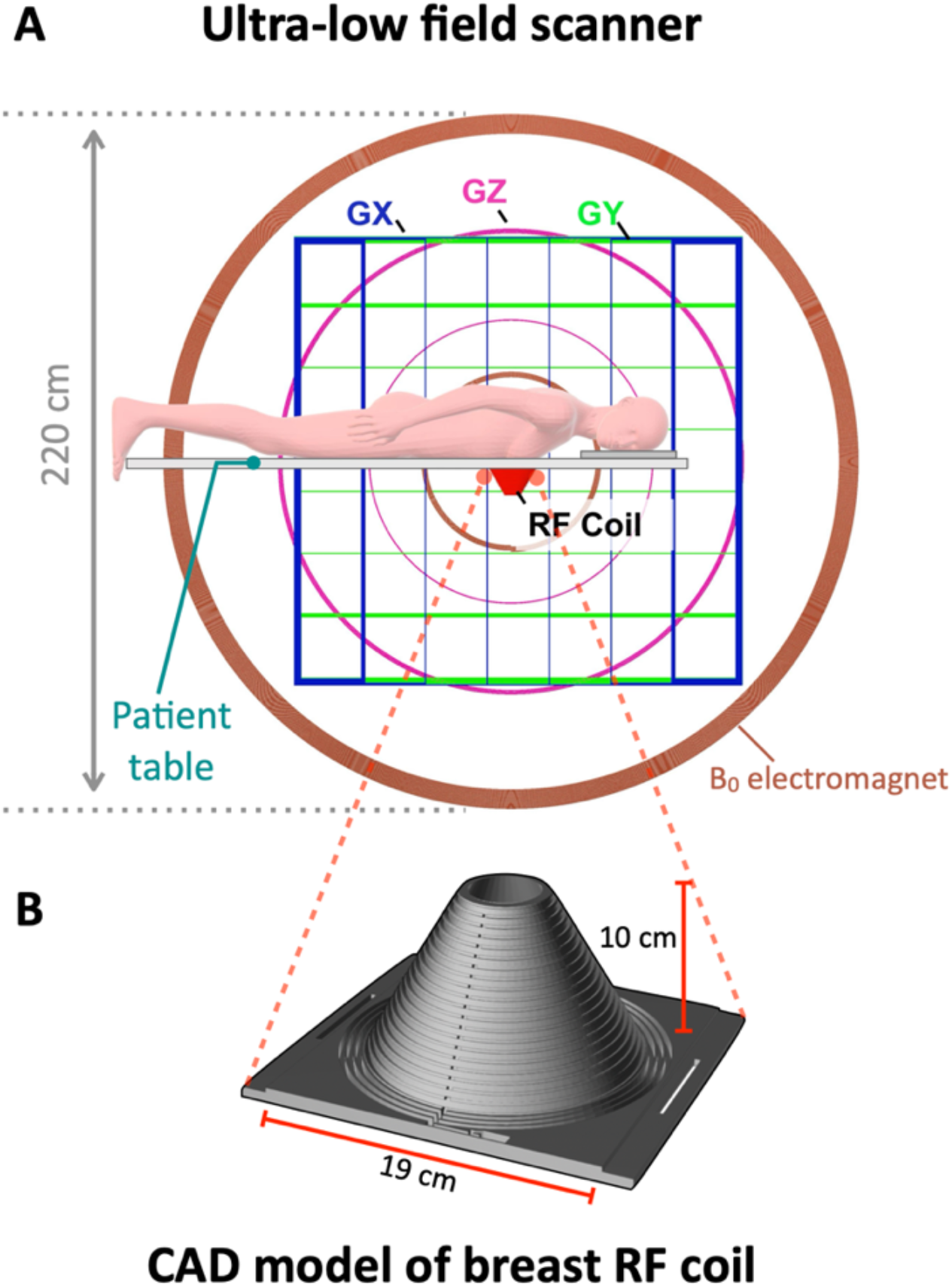
6.5 mT ULF MRI scanner configured for breast imaging. (**A**) Axial view of the ULF MRI scanner. The three axes of the gradient set are shown as Gx (in blue), Gy (in green) and Gz (in magenta), and the biplanar coils of the resistive electromagnet are shown in brown (two per side, four total). The participant lays on the patient table in the prone position with the head turned to the side. The left breast is placed in the RF coil located at the scanner isocenter. (**B**) CAD model of breast RF coil designed for breast imaging at 276.18 kHz. The dimensions of the RF coil are shown in red.

**Fig. 2.**
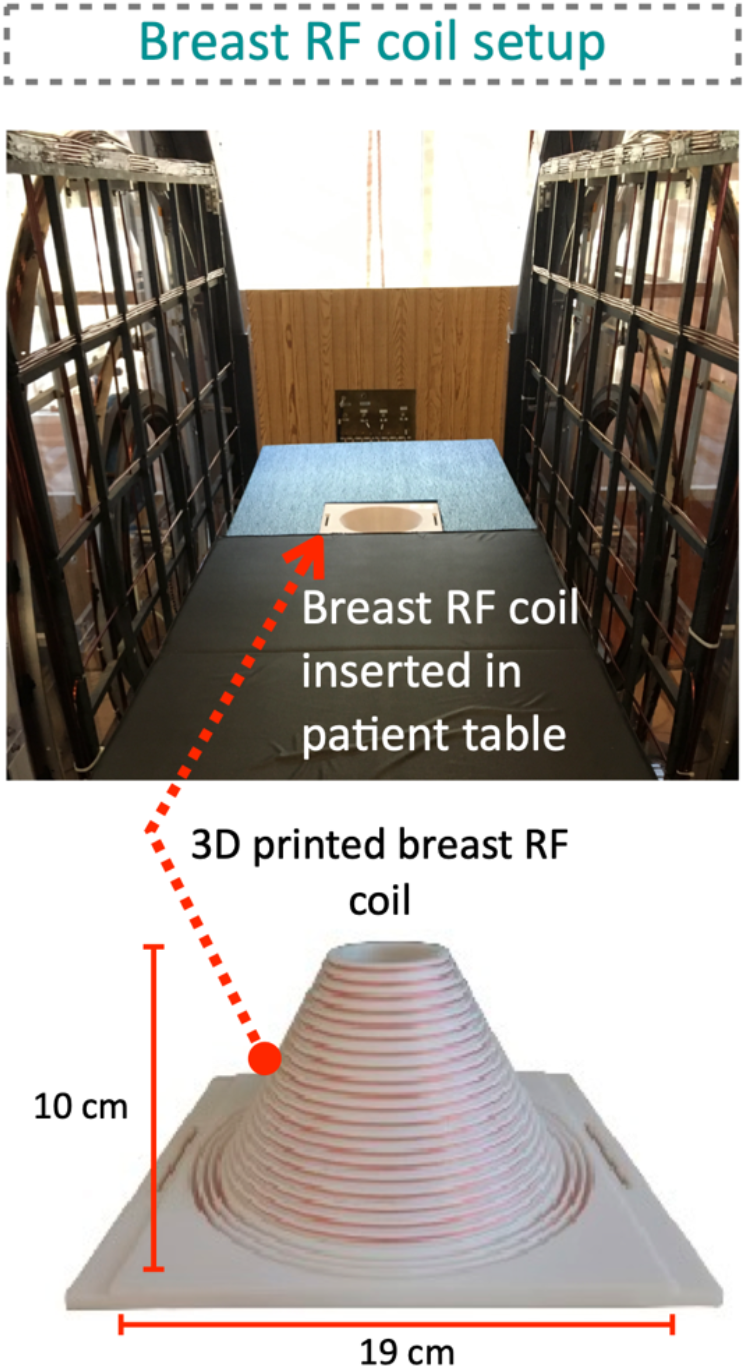
In vivo experimental setup for breast imaging at ULF. A view of the subject table with red arrow indicating the location of the 3D printed and uniformly wound breast RF coil fixed in the table below the subject. The dimensions of the RF coil are shown in red.

A close-fitting conical breast coil was designed, and the RF magnetic field generated by this coil was simulated, with the resultant field map depicted in Fig. 3A. Within the breast RF coil, the field homogeneity was quantified, revealing an inhomogeneity of ±60% in the breast volume region and a magnetic field fall-off 3 cm inside the chest wall of 30%. To evaluate the sensitivity of the coil, a homogeneous flexible phantom filled with deionized water was positioned inside the breast RF coil and scanned (Fig. 3B). The phantom imaging result, shown in Fig. 3C, reflects the sensitivity distribution across the RF coil, demonstrating high sensitivity within the coil and a marked decrease in sensitivity towards the opening of the RF coil.

**Fig. 3.**
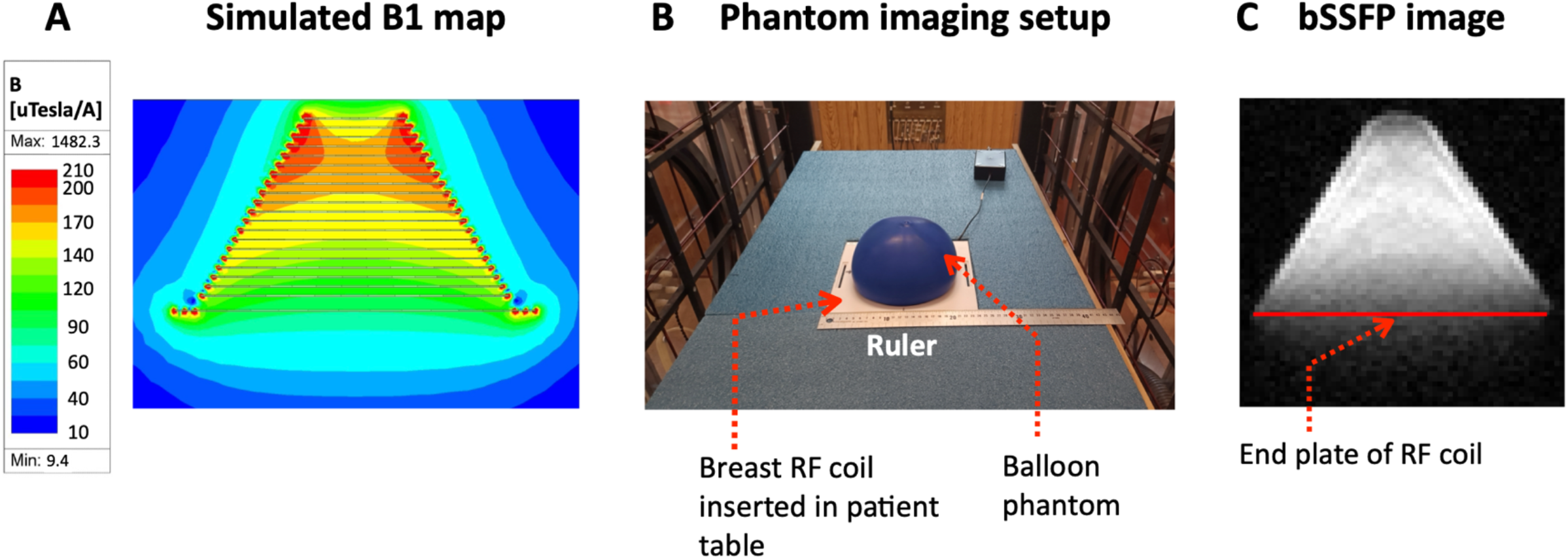
Assessment of the imaging volume of the conical-shaped breast RF coil. (**A**) The magnetic field calculation of the breast RF coil where the color bar indicates the B1 field distribution (in µT/A) across the breast volume. (**B)** A homogeneous phantom was imaged using the setup as shown, with a latex balloon (in blue) filled with deionized water placed inside the breast RF coil. (**C**) Phantom imaging scan, where a single central slice is extracted from a 21-slice 3D-bSSFP acquisition. The scan shows the signal uniformity of the RF coil. The red line indicates the end of the plate of the RF coil.

### Participant characteristics and imaging protocol

ULF MRI was used to image the left breast of 11 women (mean age, 35 years ± 13 years) in this preliminary study. All women completed the study. A 3D balanced SSFP (bSSFP) sequence was used with a voxel size of 3 mm × 3 mm × 8 mm. To accelerate the imaging process, an under-sampling factor of 70% was used, and the resulting total scan time was 21 minutes 36 seconds. The MR sequence and positioning were well tolerated. None of the images were degraded by patient motion. It is noteworthy that none of the participants experienced discomfort during the exam, and the breast fit naturally in the conical-shaped RF coil without any compression.

### ULF MRI breast imaging findings

Image sets of the entire left breast for three representative subjects are shown in Fig. 4-6. The images in Fig. 7 are single slices of these three representative subjects with the following features labeled by numbered arrows: visibility of the breast outline (indicated by 1), NAC (indicated by 2), FGT (indicated by 3), and chest wall (indicated by 4).

**Fig. 4.**
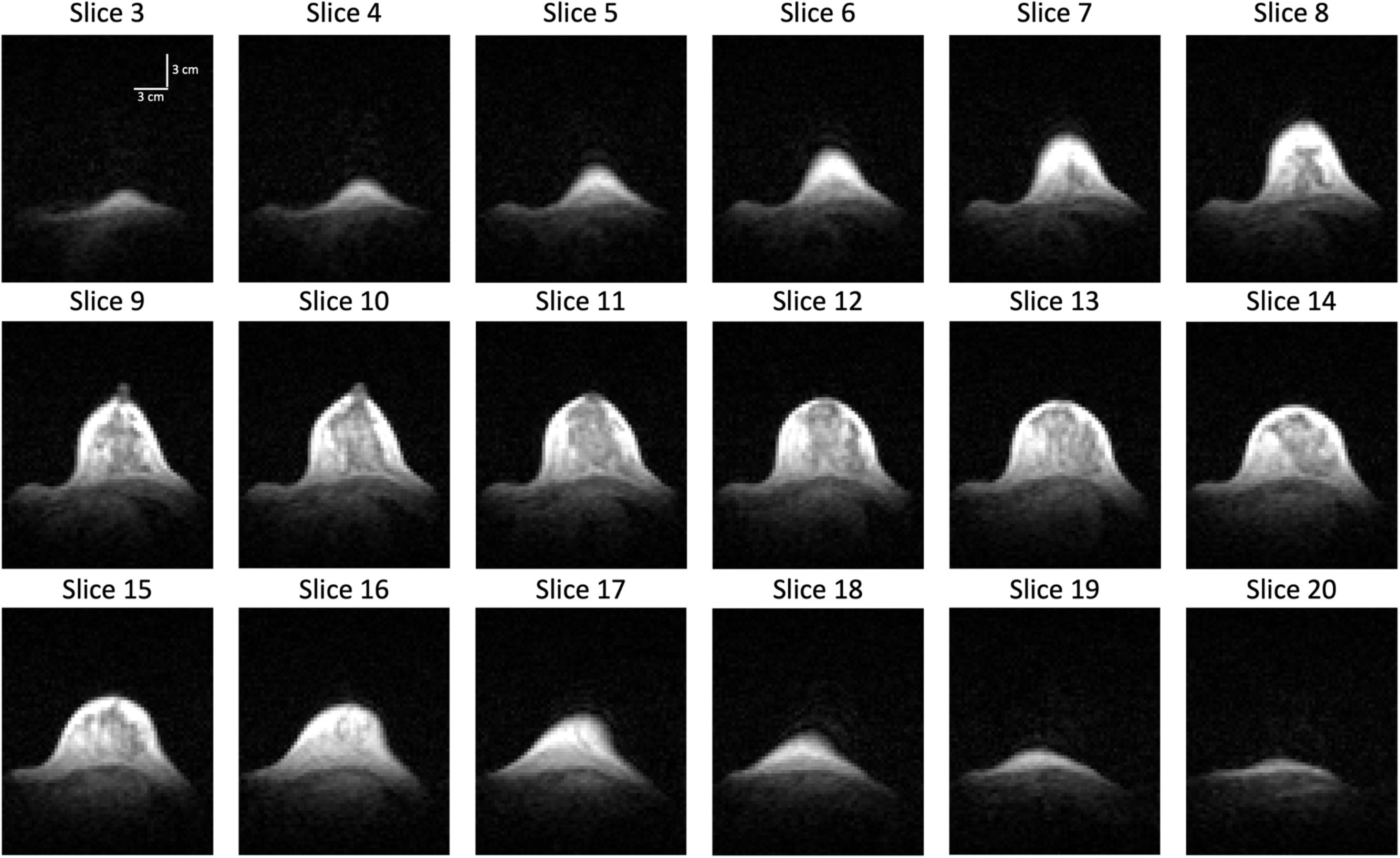
3D Ultra-low field breast MRI obtained at 6.5 mT from a healthy woman in her 30s with heterogeneous fibroglandular tissue (FGT) 18 out of 21 sequential axial bSSFP-weighted slices of the left breast are shown, and no contrast agent was administered. Data was acquired in approximately 21 minutes with a spatial resolution of 3 mm ξ 3 mm ξ 8 mm. All features are visualized in this study: breast outline, FGT, nipple areolar complex, and chest wall. Vertical and horizontal scale bars in white are 3 mm each and are shown in slice 3.

**Fig. 5.**
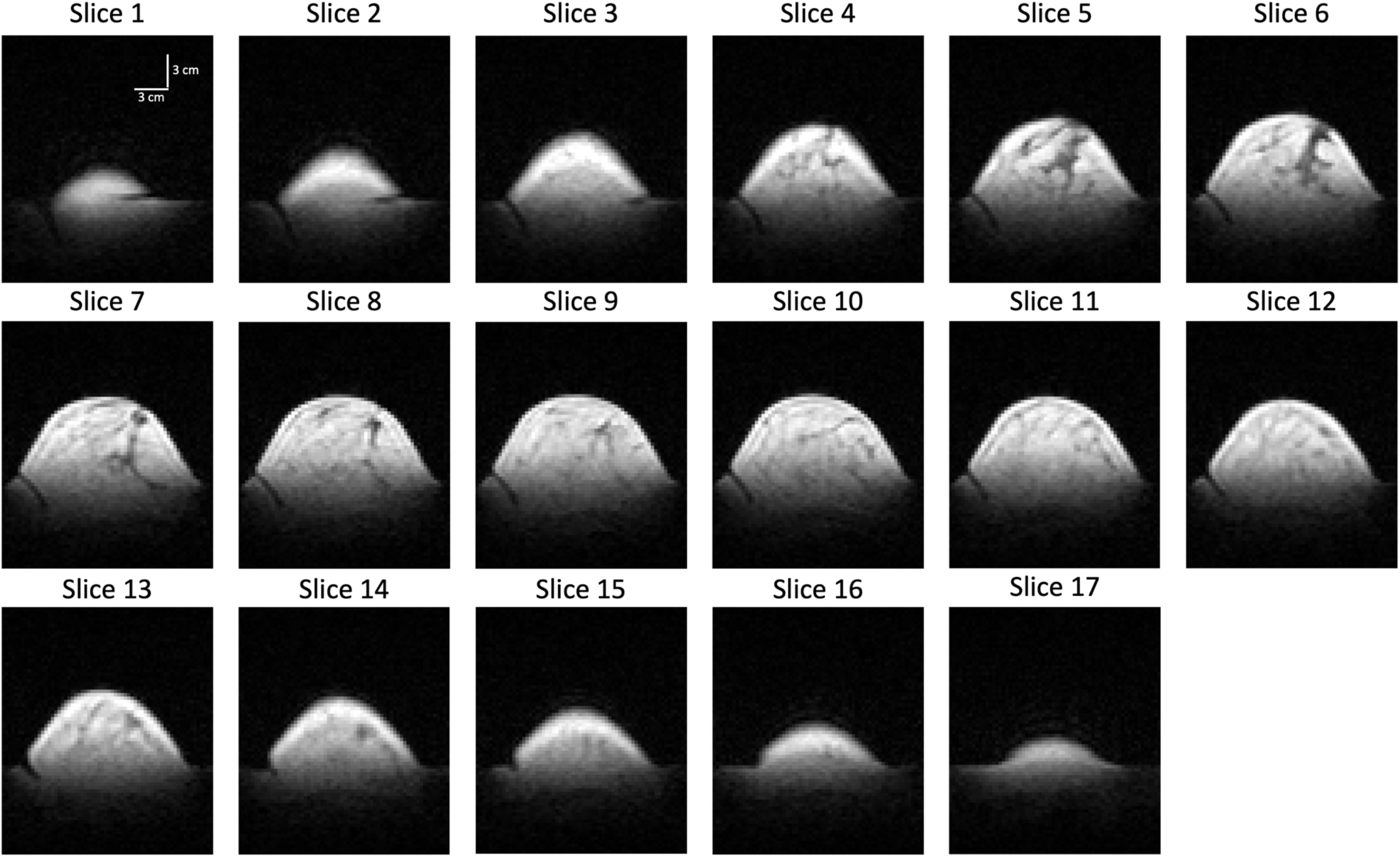
3D Ultra-low field breast MRI obtained at 6.5 mT from a healthy woman in her 30s with scattered fibroglandular tissue (FGT) 17 out of 21 representative sequential axial bSSFP-weighted slices of the left breast are shown, and no contrast agent was administered. Data was acquired in approximately 21 minutes with a spatial resolution of 3 mm ξ 3 mm ξ 8 mm. The nipple areolar complex and chest wall were not well visualized in this study. The breast outline and FGT are visualized. Vertical and horizontal scale bars in white are 3 mm each and are shown in slice 1.

**Fig. 6.**
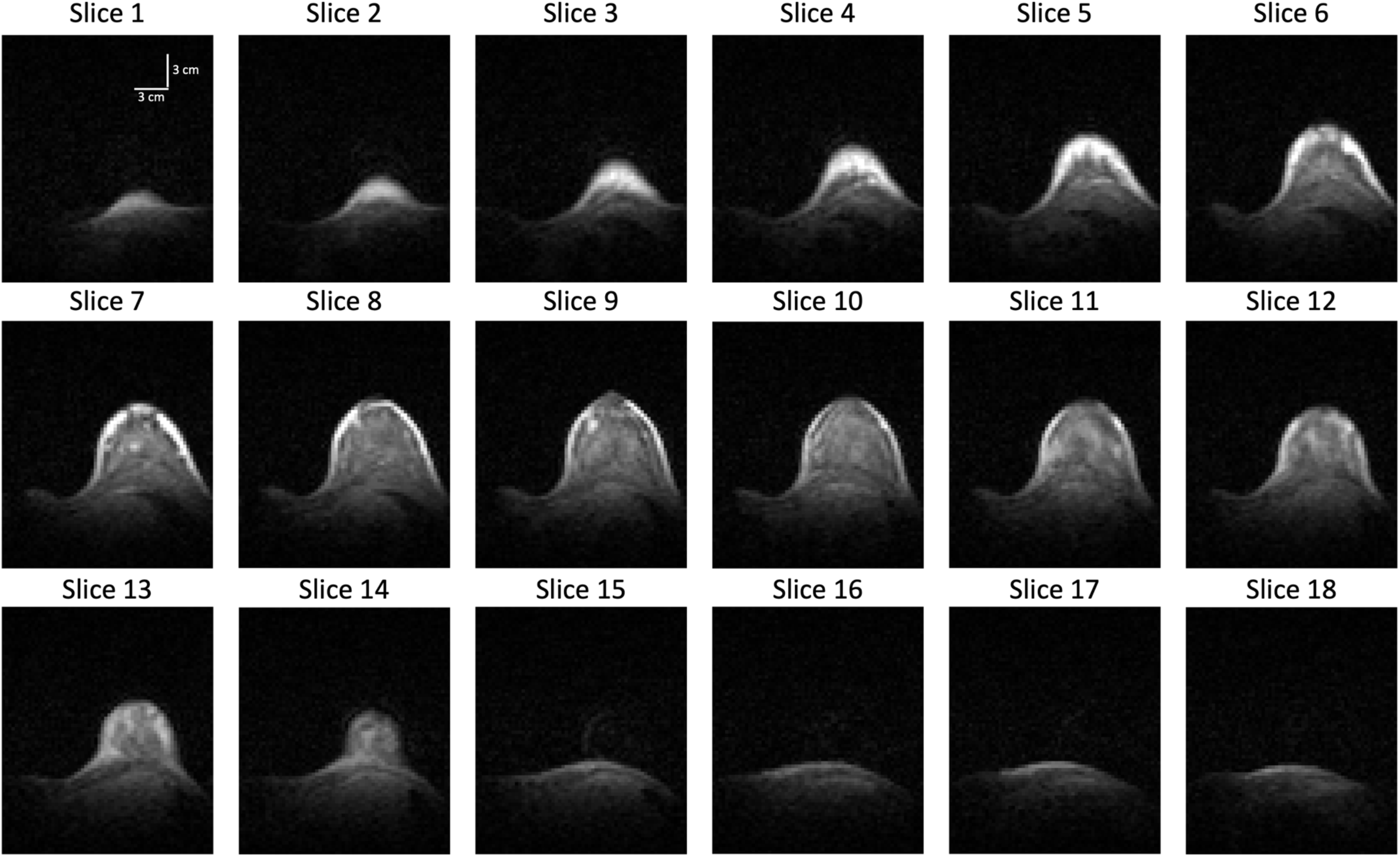
3D Ultra-low field breast MRI obtained at 6.5 mT from a healthy woman in her 30s with extreme fibroglandular tissue (FGT) 18 out of 21 representative sequential axial bSSFP-weighted images of the left breast are represented. No contrast agent was administered. Data was acquired in approximately 21 minutes with a spatial resolution of 3 mm ξ 3 mm ξ 8 mm. All features are visualized in this study: breast outline, FGT, nipple areolar complex, and chest wall. Vertical and horizontal scale bars in white are 3 mm each and are shown in slice 1.

**Fig. 7.**
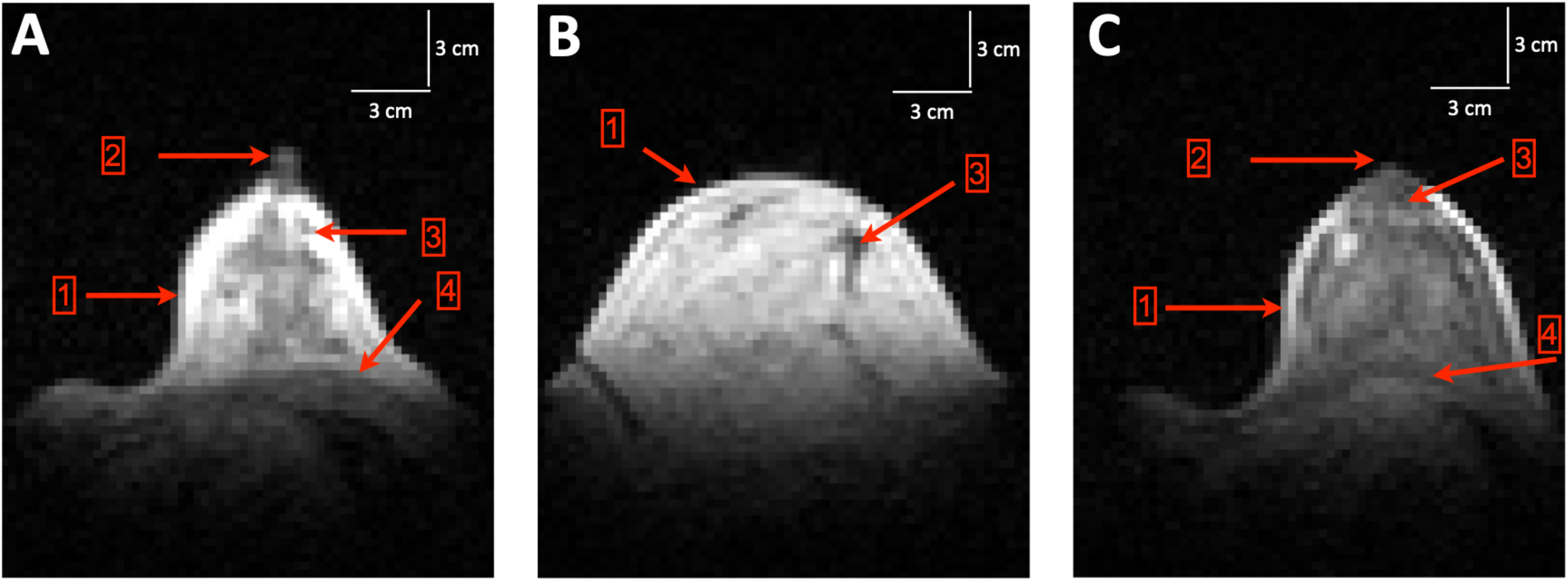
Representative axial bSSFP-weighted slices of the left breast acquired at 6.5 mT from three different subjects. (**A**) A subject with heterogeneous fibroglandular tissue (FGT) and all features visualized. (**B**) A subject with scattered FGT with the breast outline and FGT visualized. The nipple areolar complex (NAC) and chest wall were not well visualized. (**C**) A subject with extreme FGT and all features visualized. When visible, the indicated features labeled by numbered arrows are breast outline (indicated by 1), NAC (indicated by 2), FGT (indicated by 3), and chest wall (indicated by 4). Vertical and horizontal scale bars in white are 3 mm each and are shown in each slice.

Breast images from all 11 participants were evaluated by three independent board-certified breast radiologists for the purpose of categorizing breast density and assessing the visibility of essential breast tissues. Individual image scores are reported in Table 1. Breast tissue pattern was assessed using fatty, scattered FGT, heterogeneous FGT, and extreme FGT. Inter-reader reliability of breast tissue pattern was determined using Fleiss’ kappa, which resulted in a kappa value of 0.73 (95% confidence interval: 0.72 to 0.74, p<0.001), indicating substantial agreement among the readers.

**Table 1:** Qualitative assessment of imaging in each of the 11 subjects. Three breast radiologists assessed each imaging feature on a 5-point Likert scale of 1-5 (1 – not at all visible, 2 – barely visible, 3 – clearly visible but blurred, 4 – clearly visible and sharp, 5 – clearly visible and very sharp). Breast tissue pattern (density) was evaluated using F – Fatty, S – Scattered FGT, H – Heterogenous FGT, and E – Extreme FGT.

| Feature |  |  | Subject # |  |  |  |  |  |  |  |  |  |  | Statistical Analysis |  |
| --- | --- | --- | --- | --- | --- | --- | --- | --- | --- | --- | --- | --- | --- | --- | --- |
|  |  |  | Subj<br>1 | Subj<br>2 | Subj<br>3 | Subj<br>4 | Subj<br>5 | Subj<br>6 | Subj<br>7 | Subj<br>8 | Subj<br>9 | Subj<br>10 | Subj<br>11 | Fleiss Kappa test |  |
|  |  |  |  |  |  |  |  |  |  |  |  |  |  | kappa (95%<br>confidence level) | p-value |
| Breast tissue<br>pattern | Reader<br># | 1 | S | H | S | H | S | E | H | H | S | H | E | 0.73 *<br>(0.72 – 0.74) | <0.001<br>* |
|  |  | 2 | S | H | S | H | S | E | H | H | S | H | E |  |  |
|  |  | 3 | F | H | F | S | S | E | H | H | S | H | E |  |  |
| Breast outline<br>visibility | Reader<br># | 1 | 4 | 4 | 4 | 4 | 5 | 4 | 4 | 4 | 4 | 4 | 4 | 1 | - |
|  |  | 2 | 3 | 3 | 3 | 3 | 3 | 3 | 3 | 3 | 3 | 3 | 3 |  |  |
|  |  | 3 | 5 | 5 | 5 | 5 | 5 | 5 | 5 | 5 | 5 | 5 | 5 |  |  |
| Fibroglandular<br>tissue visibility | Reader<br># | 1 | 3 | 3 | 3 | 3 | 3 | 3 | 3 | 3 | 3 | 3 | 3 | 1 | - |
|  |  | 2 | 2 | 3 | 2 | 3 | 2 | 2 | 2 | 3 | 2 | 3 | 3 |  |  |
|  |  | 3 | 3 | 3 | 3 | 3 | 3 | 3 | 3 | 3 | 3 | 3 | 3 |  |  |
| Nipple Areolar<br>Complex<br>visibility | Reader<br># | 1 | 3 | 4 | 1 | 3 | 1 | 4 | 4 | 3 | 1 | 3 | 3 | 0.54<br>(0.58 – 0.6) | <0.001 |
|  |  | 2 | 2 | 3 | 1 | 2 | 2 | 3 | 3 | 2 | 1 | 3 | 3 |  |  |
|  |  | 3 | 2 | 4 | 1 | 3 | 2 | 3 | 4 | 3 | 2 | 4 | 4 |  |  |
| Chest wall<br>visibility | Reader<br># | 1 | 3 | 4 | 4 | 4 | 1 | 4 | 3 | 4 | 4 | 4 | 3 | 0.27<br>(0.26 – 0.28) | <0.2 |
|  |  | 2 | 1 | 2 | 2 | 2 | 1 | 2 | 2 | 2 | 2 | 2 | 2 |  |  |
|  |  | 3 | 3 | 4 | 4 | 4 | 3 | 3 | 4 | 3 | 4 | 3 | 2 |  |  |

Note.- Subj = Subject
\* The Fleiss kappa test on the breast tissue pattern was based on a 4-level score : 1- F, 2- S, 3- S and 4-E.
Inter-reader analysis on the 4 remaining imaging features were assessed on as a binary rating system, where scores of 2 to 5 were categorized as visible and a score of 1 as not visible.

Visibility of the following features in the breast was scored using a 5-point Likert scale (1 – not at all visible, to 5 – clearly visible and very sharp): breast outline, fibroglandular tissue (FGT) compared to intramammary adipose tissue, demarcation of the nipple areolar complex (NAC), and the chest wall, defined as visualization of the pectoralis muscle. The limited data set from this pilot study did not allow for proper training of the readers, and given the novelty of the images, the readers were not well “calibrated” to each other. For example, when evaluating the visibility of the breast outline, we find the readers were internally consistent: each reader scores all images with the same visibility (with the exception of a single case for reader 1 that received a higher score). However, each reader has assigned a different visibility score from the other readers. As a result, a binary rating system was adopted from the 5-point scale with a score of 1 remaining not at all visible and scores 2-5 as visible. Fleiss’ kappa was also used to measure the agreement regarding the visibility of essential breast tissues. In this binary framework, consensus on the visibility of the breast outline and fibroglandular (FGT) tissue was consistent (kappa = 1), whereas the nipple-areolar complex (NAC) and chest wall exhibited kappa values of 0.54 (95% confidence interval: 0.58 to 0.60, p<0.001) and 0.27 (95% confidence interval: 0.26 to 0.28, p<0.2), respectively.

## Discussion

In this preliminary study, we performed MR breast imaging at 6.5 mT on the left breast of healthy participants and were able to identify key breast features, namely breast outline, FGT, NAC, and chest wall. Eleven participants with breasts of various size were included, and images were acquired using a single bSSFP sequence lasting approximately 21 minutes. No external contrast agent was used for these studies. The results presented here encourage us to further develop ULF MRI for breast imaging, including approaches to further reduce exam time.

MRI at the low- and ultra-low magnetic field is challenging due to inherently low Boltzmann polarization and consequently low signal. Two additional consequences of MRI physics at ultra-low magnetic field are relevant to this work. First, as magnetic field decreases, tissue T1 relaxation times generally decrease, while T2 relaxation times are generally constant across fields (*32, 35*). Second, the magnetic susceptibility artifacts are significantly reduced at ultra-low field. We leverage both of these aspects to our advantage at 6.5 mT, where the efficiency of bSSFP in this regime is maximal (*34*) and enables banding-free imaging over large fields of view. In this study, the image SNR was sufficient to visualize key breast tissues.

The three expert readers had substantial agreement in their evaluation of breast tissue pattern and most key breast tissues. There were some discrepancies between the readers: specifically, the average scores of Reader 2 were 33% lower than those of Reader 1 (paired t-test, p<0.001) and 28% lower than Reader 3 (paired t-test, p<0.001), whereas the scores of Reader 3 were 7.01% higher than those of Reader 1 (paired t-test, p<0.03). Also, the readers had some disagreement on the visibility of the NAC and chest wall. We attribute this to two factors: lack of training and lack of experience with ULF MRI. The limited data set did not allow for proper training of the readers, and instead only the evaluation criterion were discussed. Additionally, there is a lack of calibration across the readers, given that these are their first experiences with ULF MRI images. Conversely, if these readers were examining clinical breast MRI scans, there would be an implicit calibration, since the readers have all examined many clinical breast MRI scans, over a long period of time (13 years, 3 years and 9 years, respectively).

The NAC and chest wall were not always visible. The absence of NAC on certain scans can be due to either the slice thickness and positioning of the breast or the normal variations in human anatomy, which include flat or inverted nipples. The chest wall was not always visible, primarily in participants with a larger breast. This is a limitation of the coil design. Since the imaging depth of the RF coil is approximately 3 cm from the end plate of the RF coil, the chest wall was not fully captured in participants with larger breast sizes.

Our current methods have some limitations. In addition to the lack of visualization in the chest wall, our preliminary study did not image the axilla, a potential site of breast cancers and nodal disease. Also, the image resolution used here falls short of the clinical requirements for breast cancer screening where a target resolution of 2 mm× 2 mm × 5 mm is needed to identify small tumors. Ideally, both breasts and axilla could be imaged simultaneously at the target resolution in a scan time of ten minutes or less. To decrease the total exam time, the use of RF coils capable of imaging both breasts simultaneously with a field of view that includes the axilla and chest wall can be developed. Our relatively simple low-cost coil design allows the construction of breast coils in a variety of sizes to maximize the filling factor and thus the SNR for a given subject (*36*).

The 6.5 mT ultra-low field magnetic resonance imaging system used here is a configurable test bed system developed in our laboratory to perform preliminary research and refine sequences and techniques for breast cancer imaging. To be considered for clinical use, it is necessary to increase the SNR, as improved SNR can be used to attain increased resolution, decreased scan time, or both. Although the results shown here were acquired at 6.5 mT, operation at even moderately higher magnetic field (B_0_) will have a big impact on increasing the attainable resolution and decreasing the scan time for this application. A factor of 3 increase in magnetic field to a nominal 20 mT will result in a factor of 5 increase in SNR, as SNR is proportional to B_0_^3/2^ (*37*). This could allow us to obtain images 25x faster for the same SNR. Further increases in SNR from an increase in field strength to approximately 65 mT (10x higher than the studies presented here) could maintain the mobility and low-cost of a low-field system but with a very significant reduction of imaging time and an increase in spatial resolution.

We note that the absolute chemical shift between fat and water decreases with decreasing field, making conventional water suppression techniques more challenging. Previous work using NMR and NMR dispersion techniques observe that the T1 of adipose tissue in the breast does not change with field strength, while the T1 of fibroglandular tissues do change with field strength (*32, 33*). Thus, it may be possible to make a fat suppression technique that takes advantage of the T1 dispersion differences.

Contrast agents are typically used to increase the contrast between a tissue of interest and the surrounding tissue, and clinical breast MRI requires the use of contrast agents to identify breast tumors (*38–40*). However, there is concern about the long-term effects of repeated administration of MRI contrast agents such as gadolinium (*41*). At low magnetic fields, however, gadolinium-based contrast agents do not improve the contrast of the image, in part because gadolinium is not magnetically saturated at low magnetic fields, and thus does not increase the brightness of the image. Recent work highlights the possibilities of iron-oxide nanoparticles and SPIONS for use at low magnetic fields (*42, 43*). A possible benefit of iron-oxide based agents is their biocompatibility, and preliminary in vivo studies used ferumoxytol, an FDA-approved SPION-based treatment of iron deficiency anemia (*44, 45*). Contrast agents were not used in this preliminary study.

With this perspective on low-field MRI physics, these initial results encourage us to envision many possibilities for non-contrast, low-field, breast MRI. A purpose-built system could be used in many possible imaging orientations including prone as is current practice, but also extending to supine in the surgical orientation, sitting, or standing. Different magnet designs can be considered to enable portability, low-cost, and integration in a surgical suite or other locations where the magnetic fields of typical clinical systems (1.5 T and 3 T) prohibit safe imaging. ULF MRI systems can increase access, and the absence of gadolinium-based intravenous contrast agents and enclosed spaces may increase use of screening breast MRI.

In conclusion, we have demonstrated the feasibility of ultra-low field magnetic breast imaging without the use of contrast agents or compression. This approach may provide a new option for breast cancer screening and diagnosis in the future.

## Materials and Methods

### Study Design and Participants

This prospective pilot study was performed from March 2023 to May 2023 and granted institution review board approval from the Office for Human Research Studies (protocol 21-579) at the Dana-Farber/Harvard Cancer Center. Written informed consent was obtained from each participant.

A total of 11 healthy female participants were enrolled (mean age, 35 years ± 13 years). Exclusion criteria were: pregnancy, breastfeeding, or inability to undergo MRI due to presence of an implanted or external MRI unsafe device or MR conditional device not meeting the conditions required for the scan. Participants had to be older than 20 and younger than 80 years old. The study also excluded individuals directly supervised by study investigators.

### Imaging System

Imaging was performed on a custom-built electromagnetic MRI scanner, shown in Fig. 1, and previously described (*34*). The scanner operates at a main field strength of 6.5 mT (Larmor frequency of 276.18 kHz). The shimmed magnetic field inhomogeneity measured over a 20 cm spherical region at isocenter is less than 10 Hz. Imaging gradients are produced by a biplanar gradient set capable of producing linear gradients of up to 1 mT/m in all three axes.

For this study, the imaging bed was modified from its previous configuration for neuroimaging (*34*) to a breast imaging setup where the breast and breast RF coil are located at the isocenter of the scanner. Figure 2 illustrates the imaging bed and dedicated RF coil designed to image a single breast. In order to achieve a good filling factor and thus a high SNR (*36*), a close-fitting conical breast RF coil was designed. To evaluate RF coil homogeneity, the magnetic field was calculated using the Finite-Element-Method simulation (Ansys Maxwell, 2021, Ansys, Canonsburg, PA, USA). The simulated magnetic field was used to assess the field homogeneity within the breast volume and to determine the magnetic field fall-off beyond the physical end of the coil. The uniformity of the breast imaging region was also assessed using a homogeneous flexible phantom consisting of a latex balloon filled with deionized water. This MR phantom was placed inside the breast RF coil, and as seen in Fig. 3B, it occupied the entire imaging region-of-interest. The imaging protocol used to scan the MR phantom was the same as that of participant scanning protocol.

The decision in favor of this type of coil shape was mainly based on promising study results at higher field strengths (*46*). The conical RF coil was also adapted in size to enable imaging of larger breasts, based on the reported common female breast sizes in the US (*47*). The coil height is 10 cm; its diameter at the base is 19 cm; and its diameter at the peak is 4 cm (*48, 49*). The RF coil is uniformly wound on a conical supporting structure. This coil design is capable of imaging the whole breast and the chest wall to a depth of approximately 3 cm. Using this breast coil, the left breast of all participants was imaged with participants in the prone position, as schematically illustrated in Fig. 1A. Photographs of the actual *in vivo* experimental setup can be requested from the corresponding author.

### MRI acquisition

A 3D balanced SSFP (bSSFP) sequence was used with a flip angle of 70 degrees, TE (echo time)/TR (repetition time) of 13 ms/26 ms, a matrix size of 64 × 72 × 21, 50 averages, and a voxel size of 3 mm × 3 mm × 8 mm. To accelerate the imaging process, an under-sampling factor of 70% was used. The total scan time was 21 minutes 36 seconds. No contrast agents were used. Given the scan duration, the study was limited to imaging one breast, and the left breast was imaged in all participants for consistency.

Images were reconstructed in MATLAB (Natick, MA, USA) using inverse fast Fourier transform (IFFT) with the under-sampled region zero-filled in k-space. Images were converted into DICOM format using the MATLAB function dicomwrite.

The MR images of all participants were reviewed by three board-certified breast radiologists (M.A.S., L.R.L and J.C.VC) with 13, 9 and 3 years of experience reading breast MRI. The readers reviewed the evaluation criteria; however, due to the limited data of this pilot study, no additional images were used to train the readers. Images were viewed in DICOM format using 3D Slicer (*50*). The visibility of the following features in the breast was assessed: visibility of the breast outline, visibility of the fibroglandular tissue (FGT) compared to intramammary adipose tissue, demarcation of the nipple areolar complex (NAC), and visualization of the pectoralis muscle (chest wall). Visibility of these features was assessed using a 5-point Likert scale (1 – not at all visible, 2 – barely visible, 3 – clearly visible but blurred, 4 – clearly visible and sharp, 5 – clearly visible and very sharp). Breast tissue pattern (density) was assessed using four categories: fatty, scattered FGT, heterogeneous FGT, and extreme FGT. Images were also evaluated for motion artifacts.

### Statistical Analysis

Inter-reader agreement was assessed by computing Fleiss’ kappa among three reader’s feature visibility assessments. Due to the novelty of these images, i.e., they were new to all readers, and the limited data set, which did not allow for proper training of the readers, the readers were not “calibrated” to each other, as they are when reading clinical MRI. As a result, the 5-point scale was revised to a binary scale to assess whether or not a feature was visible (1 – not at all visible, 2 or greater – visible). All statistical analyses were performed using IBM SPSS Statistics for Windows, version 26.0. (IBM Corp., Armonk, NY, USA).

## Data Availability

All data generated or analyzed during the study are available in the main text.

## Acknowledgments

The authors would like to thank Darrah Bowden for her invaluable assistance and perspective on the breast imaging configuration. MSR dedicates this work to the memory of Christina Pfeifer Mattig.

## Author contributions

Conceptualization: KEK, MSR

Methodology: SS, NK, FKL, MAS, TPPH, SEO

Investigation: SS, NK, FKL, MAS, LRL, JVCV, TPPH, MSR

Visualization: SS, NK, MAS, JVCV, SEO, KEK, MSR

Supervision: SY, TRB, KEK, MAS, MSR

Writing—original draft: SS, NK, KEK, MSR

Writing—review & editing: SS, NK, FKL, MAS, LRL, JVCV, TPPH, SEO, SY, TRB, KEK, MSR

## Data and materials availability

All data generated or analyzed during the study are available in the main text.

## NIST Disclaimer

Certain commercial equipment, instruments, or materials are identified in this paper in order to specify the experimental procedure adequately. Such identification is not intended to imply recommendation or endorsement by NIST, nor is it intended to imply that the materials or equipment identified are necessarily the best available for the purpose.

